# Comparing COVID-19 and influenza presentation and trajectory

**DOI:** 10.1101/2020.11.19.20235077

**Authors:** Anat Reiner Benaim, Jonathan Aryeh Sobel, Ronit Almog, Snir Lugassy, Tsviel Ben Shabbat, Alistair Johnson, Danny Eytan, Joachim A. Behar

**Affiliations:** Rambam Health Care Campus, Haifa, Israel; Faculty of Biomedical Engineering, Technion, Israel Institute of Technology, Haifa, Israel; MIT Institute for Medical Engineering and Science, Massachusetts Institute of Technology, Cambridge, Massachusetts, USA

**Keywords:** COVID-19, influenza, SARI, biomarkers

## Abstract

**Background:** COVID-19 is a newly recognized illness with a predominantly respiratory presentation. It is important to characterize the differences in disease presentation and trajectory between COVID-19 patients and other patients with common respiratory illnesses. These differences can enhance knowledge of pathogenesis and help in guiding treatment.

**Methods:** Data from electronic medical records were obtained from individuals admitted with respiratory illnesses to Rambam Health Care Campus, Haifa, Israel, between October 1st, 2014 and October 1st, 2020. Four groups of patients were defined: COVID-19 (693), influenza (1,612), severe acute respiratory infection (SARI) (2,292) and Others (4,054). The variable analyzed include demographics (7), vital signs (8), lab tests (38),and comorbidities (15) from a total of 8,651 hospitalized adult patients. Statistical analysis was performed on biomarkers measured at admission and for their disease trajectory in the first 48 hours of hospitalization, and on comorobidity prevalence.

**Results:** COVID-19 patients were overall younger in age and had higher body mass index, compared to influenza and SARI. Comorbidity burden was lower in the COVID-19 group compared to influenza and SARI. Severely- and moderately-ill COVID-19 patients older than 65 years of age suffered higher rate of in-hospital mortality compared to hospitalized influenza patients. At admission, white blood cells and neutrophils were lower among COVID-19 patients compared to influenza and SARI patients, while pulse rate and lymphoctye percentage were higher. Trajectories of variables during the first two days of hospitalization revealed that white blood count, neutrophils percentage and glucose in blood increased among COVID-19 patients, while decreasing among other patients.

**Conclusions:** The intrinsic virulence of COVID-19 appeared higher than influenza. In addition, several critical functions, such as immune response, coagulation, heart and respiratory function and metabolism were uniquely affected by COVID-19.

## Introduction

Severe acute respiratory syndrome coronavirus 2 (SARS-CoV-2), is the the virus underlying COVID-19, a newly recognized illness that initially spread throughout Wuhan (Hubei province), and from there, to other provinces in China and then across the globe. As of October 15th 2020, over 39,000,000 infections and over 1,100,000 casualties have been linked to SARS-CoV-2. The clinical spectrum of SARS-CoV-2-associated pneumonia ranges from mild to to life-threatening(1, 2). Several studies have described general epidemiological findings, clinical presentation and clinical outcomes of SARS-CoV-2 pneumonia, and identified mortality risk factors(3–9). The need for detailed information on the clinical characteristics of hospitalized patients with COVID-19 and their clinical course is essential to achieve a thorough understanding of the disease development and progression. Moreover, the exact differences between the clinical presentation and illness trajectory of COVID-19 versus other respiratory (viral) infections remain illusive. Investigating the clinical features of influenza like illness (ILI) is of paramount importance to identify COVID-19 specificities. This work questioned the virulence of COVID-19 as compared to seasonal influenza and SARI. Recognizing the characteristics discriminating COVID-19 from influenza, will be critical to support the management of the current pandemic. There are limited works that have pursued the pathophysiological differences between ILIs and COVID-19, and have focused solely on the H1N1 influenza strain (10–13). They investigated symptoms, comorbidities, laboratory examinations, treatments and scans in relatively small cohorts of patients and consequently had limited statistical power. Hence, the relative virulence of COVID-19 versus influenza has been and remains under debate. Discriminating biomarkers at clinical presentation and characteristics of the trajectory of COVID-19 versus influenza are poorly characterized.

To address this knowledge gap, this retrospective analysis reviewed the electronic medical records (EMR) from the Rambam Health Care Campus, located in Haifa, Israel. Demographics, comorbidities, vital signs and laboratory tests were analyzed to identify features that can potentially discriminate between COVID-19, influenza and SARI at the time of admission to the hospital, as well as their trajectory during the first 48 hours after admission.

## Methods

### Data Source

A unique cloud-based database, named COV19, was created based on the model of MIMIC III (14). The database contained detailed de-identified clinical information. Specific views (tables) that contained multiple variables related to a given type of medical data, were created.

Ethical approval for this research was provided by the local institutional review board (IRB; #0141-20). The de-identified datasets were uploaded to a Microsoft Azure cloud server, which also offers data analysis, visualization and querying tools. The description of the tables and variables included in COV19 is available on the online resource site (https://cov19-resource.com/). Future access to the cloud can be given to interested researchers, subject to hospital IRB approval.

### Study population

This single center retrospective observational cohort study uses EMR data from Rambam Health Care Campus, a 1000-bed tertiary academic hospital in Northern Israel, during which the pandemic opened five dedicated COVID-19 departments. The hospital EMR database was queried for hospitalized adult (age 18 and above) cases admitted for COVID-19, influenza or SARI, or tested for COVID-19 during hospitalization (with either a positive or negative result), from October 1st, 2014 until October 1st, 2020.

#### Disease groups

The disease groups were defined as follows: **COVID-19**: At least one positive reverse transcription polymerase chain reaction (RT-PCR) test for SARS-CoV-2 in nasopharyngeal swab. Most COVID-19 cases were positive within a week before admission to the hospital or at admission, and very few of them were diagnosed a few days after admission. COVID-19 were also tested for Influenza. **Influenza**: tested positive for influenza A or B virus by RT-PCR test and tested negative for COVID-19, or tested positive for influenza A or B virus by RT-PCR test and admission date prior to COVID-19 emergence in Israel, on Februrary 23, 2020. **SARI only**: Physician report in the EMR that matches World Health Organization (WHO) SARI case definition (15) i.e., an acute respiratory infection with history of fever or measured fever of 38 °*C*, and cough with onset within the last 10 days, hospitalization, and no positive test for COVID-19 or influenza (either negative or not tested). **Others**: Tested negative for COVID-19 and not classified into influenza or SARI groups.

40% of the SARI cases were tested for Influenza, when indicated by the physician for differential diagnosis or for the purpose of surveillance during the influenza season. As the cohort is defined by inclusion criteria as all patients tested for COVID-19, or either testing positive for influenza or having a SARI diagnosis, the “others” group is a control group inevitably generated by those with negative result for COVID-19, that are also negative for Influenza and SARI. Thus this group contains zero cases prior to February 23, 2020, the date of COVID-19 emergence in Israel. After this date, which approaches the end of the Influenza season, only 5% of the patients in “others” group were tested for Influenza. Tests for COVID-19 after its emergence were performed for 80% of the SARI patients, and by inclusion criteria, for all patients in “others”.

#### COVID-19 Severity

Following existing guidelines within the context of COVID-19 (16–18), patients were defined as moderately ill if they were diagnosed for COVID-19 pneumonia clinically or by x-ray. Patients were defined as severely ill if either their breaths number per minute was larger than 30, their unsupported oxygen saturation was of 93% or lower, their PF ratio was below 300, or were critically ill. Critically ill patients were defined as those who either went through mechanical ventilation support (invasive or non-invasive), were hospitalized in an intensive care unit, or were administered vasopressor medications (noradrenaline and vasopressin) or inotropic medications (dopamine, dobutamine, milrinone and adrenaline).

#### Collected Data

Demographic information such as age, sex, ethnic group (Jewish or Arabs, which included Druze and Muslims),weight, body mass index (BMI), length of hospitalization and mortality rates were collected. In addition, comorbidities, vital signs including fever, respiratory variables (breaths count per minutes, oxygen saturation) blood pressure and tests results such as metabolic profiles, complete blood count and coagulation tests were collected. Comorbidities were defined by ICD-9 codes as detailed in supplementary Table S1. A thorough analysis (see “Statistical Analysis” Section) of comorbidities, demographics and mortality rate was performed to characterise predispositions and the severity and case fatality of each disease. Moreover, lab tests and vital signs were screened at admission and for the first two days of hospitalization in order to identify distinct signature or putative biomarkers of COVID-19, influenza and SARI.

#### Statistical Analysis

Demographic variables, comorbidity rate, vital signs and lab tests were compared between disease groups at admission using the Chi-squared test or Fisher’s exact test for categorical variables, and analysis of variance or Kruskal-Wallis test for continuous variables. The p-values across all tests were corrected to control the false discovery rate (FDR) criterion (19). Mortality rates were compared before and after excluding patients at mild severity level. Medians and inter-quartile range (IQR) were used to describe the continuous variables. In addition, standardized scores were calculated for scale unification across variables, using the median for centralization and the median absolute deviation (MAD) for rescaling, thereby allowing their representation in a comparative heatmap. Adjustments for confounders were performed using generalized linear models. Age-adjusted COVID-19 odds ratios for each comorbidity were calculated using multivariate logistic regression, excluding mild cases, to eliminate severity bias.

The trajectory over time was compared for each numeric measure, using a non-parametric repeated measure model for a factorial design (20). In the first stage, for each measure, we used the interaction effect in the model to test for difference in time trend between the disease groups. For variables that showed a significant effect, we conducted post-hoc pairwise tests between the groups. The p-values over all tests across the two stages were corrected using a hierarchical FDR controlling procedure (21). Three time intervals were defined to follow trends during hospitalization: 0-6, 6-24, and 24-48 hours from admission. Effect size was defined by the difference in slopes across time between each pair of compared disease groups. The larger slope difference among the slopes obtained for (0-6, 6-24) and (6-24, 24-48) time gaps was selected as the effect size for each pairwise comparison. The R software (22) was used for statistical analysis, including the R package nparLD (23) for applying the non-parametric model. A 0.05 threshold was used to determine significance.

## Results

### Database

A total of 9,670 hospitalizations at Rambam Health Care Campus met the initial inclusion criteria between October 1st, 2014 and October 1st, 2020 (Figure 1). Excluded were 33 non-critical COVID-19 cases admitted before April 1, 2020, a period during which all positive cases were systematically hospitalized, including very mild and asymptomatic cases, and 23 COVID-19 cases admitted for reasons unrelated to COVID-19 (e.g., women at labor, traffic accident injury). In order to remove bias and correlations due to repeated per-patient hospitalizations 963 cases, mainly non-COVID-19 hospitalization, with fewer than 30 days between consecutive hospitalization, were excluded. The remaining 8,651 cases were classified into the four disease groups. This included 693 COVID-19 patients, of whom 127 (18.3%) were classified as critical. A total of 68 variables were evaluated: demographics (7), vital signs (8), lab tests (38) and comorbidities (15).

**Fig. 1.**
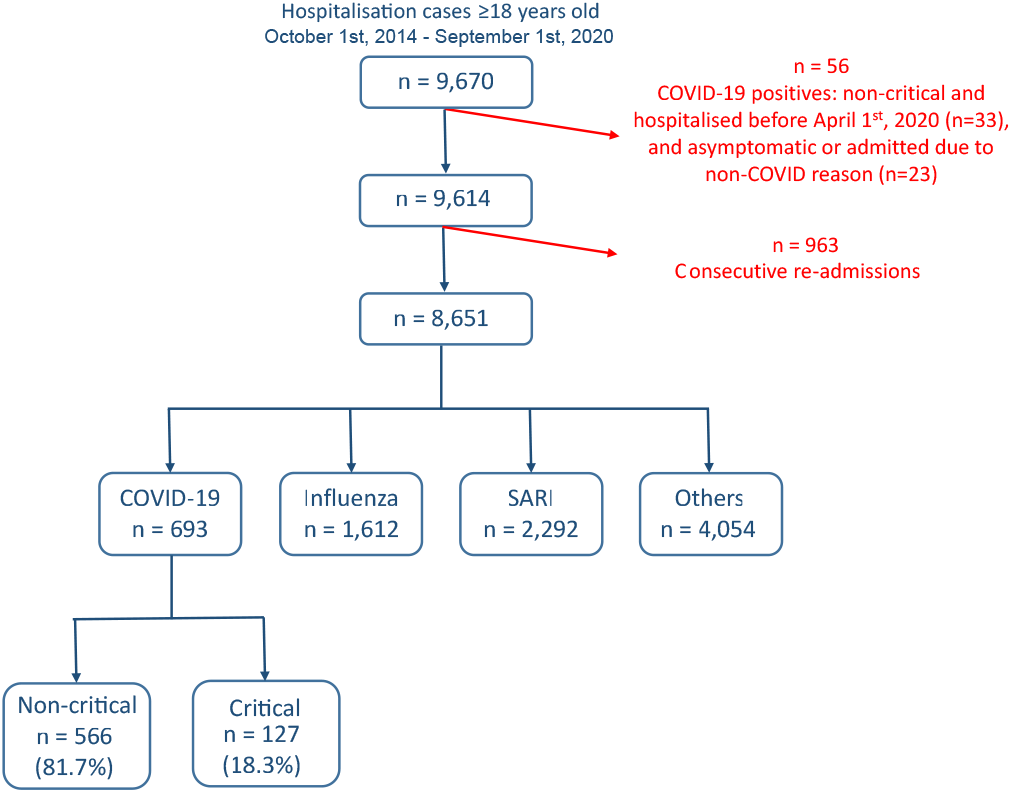
Cohort selection and criteria for exclusion. Data from a total of 9,670 admitted cases were extracted from the Rambam Health Care Campus electronic medical records system.

### Demographics

As shown in Table 1, COVID-19 patients (median age 59,8 ± 29,7 IQR *years*) were younger compared to influenza patients (median age 70,5 ± 21,8 IQR *years*) and SARI patients (median age 70,5 ± 22,9 IQR *years*). Patients between 18 and 44 years of age made up 25% of the COVID-19, compared to 13%-14% among influenza and SARI patients. In contrast 24% of the COVID-19 patients were 75 years or older, compared to 40% among influenza and SARI patients. The sex distribution among COVID-19 patients reflected a small preference towards males (52%), compared to a more significant preference towards males among SARI patients (58,6%), and no preference among influenza patients (50%). The proportion of Arabs among the COVID-19 patients (40%) was larger compared to influenza and SARI patients (25% and 23,5%, respectively), independently of disease severity. Duration of hospitalization was shorter for COVID-19 patients (median 4 ± 7 IQR *days*) compared to influenza and SARI patients (respectively median 5± 5 IQR and median 5± 6 IQR *days*). However, for moderate to severe COVID-19 patients, a longer duration of hospitalization was observed (median 6± 7,75 IQR *days*) as compared to influenza and SARI. BMI was higher for COVID-19 patients (median 28,7± IQR 7,2 *kg/m*^2^) compared to influenza (median 27,8 ± IQR 7,3 *kg/m*^2^) and SARI (median 26,6 ± 7,4 IQR *kg/m*^2^), with 35% of the COVID-19 patients being obese (BMI>30 *kg/m*^2^), compared to 30% of the influenza patients and 24% of the SARI patients. Moreover, 48% of moderate to severe COVID-19 patients under 65 years of age with no comorbidities were obese, compared to 26% of influenza patients and 14% of SARI patients. After adjustment for age, BMI among moderate to severe COVID-19 patients was found larger by 1,7 *kg/m*^2^ compared to influenza patients, and by 2,9 *kg/m*^2^ compared to SARI patients.

**Table 1.** Patient characteristics, length of stay and mortality

|  |  | COVID-19<br>(n=693) | Influenza<br>(n=1612) | SARI<br>(n=2292) | others<br>(n=4054) | FDR corrected p-value<br>(group difference) |
| --- | --- | --- | --- | --- | --- | --- |
| Age (*years*) | median (IQR) | 59,8 (29,7) | 70,5 (21,8) | 70,5 (22,9) | 66,3 (36,9) | 2,04E-46 |
| Age group | 18-44 | 176 (25,4%) | 225 (14,0%) | 296 (12,9%) | 1040 (25,7%) | 8,24E-58 |
|  | 45-54 | 101 (14,6%) | 103 (6,4%) | 168 (7,3%) | 334 (8,2%) |  |
|  | 55-64 | 126 (18,2%) | 266 (16,5%) | 329 (14,4%) | 487 (12,0%) |  |
|  | 65-74 | 124 (17,9%) | 358 (22,2%) | 546 (23,8%) | 808 (19,9%) |  |
|  | 75+ | 166 (24,0%) | 660 (40,9%) | 953 (41,6%) | 1385 (34,2%) |  |
| Gender | Male | 361 (52,1%) | 806 (50,0%) | 1344 (58,6%) | 1870 (46,1%) | 1,43E-19 |
|  | Female | 332 (47,9%) | 806 (50,0%) | 948 (41,4%) | 2184 (53,9%) |  |
| Ethnic group | Arab | 273 (39,4%) | 401 (24,9%) | 539 (23,5%) | 1000 (24,7%) | 3,05E-16 |
|  | Jewish | 420 (60,6%) | 1210 (75,1%) | 1753 (76,5%) | 3052 (75,3%) |  |
| Length of stay (*days*) | median (IQR) | 4 (7) | 5 (5) | 5 (6) | 6 (8) | 1,17E-25 |
| Weight (*kg*) | median (IQR) | 80 (22) | 75 (21) | 72 (25) | 75 (23,1) | 9,16E-14 |
| BMI (*kg/m<sup>2</sup>*) | median (IQR) | 28,7 (7,2) | 27,8 (7,3) | 26,6 (7,4) | 27,5 (7,4) | 7,26E-16 |
| BMI group | <=20 | 20 (3,5%) | 89 (8,0%) | 175 (12,7%) | 215 (7,0%) | 1,83E-16 |
|  | 20-25 | 140 (24,4%) | 301 (27,1%) | 451 (32,7%) | 968 (31,6%) |  |
|  | 25-30 | 211 (36,8%) | 382 (34,4%) | 418 (30,3%) | 1036 (33,8%) |  |
|  | 30< | 202 (35,3%) | 339 (30,5%) | 334 (24,2%) | 845 (27,6%) |  |
| Death (in hospital) | overall | 50 (8,0%) | 131 (8,1%) | 308 (13,5%) | 331 (8,6%) | 3,21E-10 |
| Death by age | 18-44 | 0 (0%) | 4 (1,8%) | 17 (5,7%) | 19 (2%) | 5,09E-04 |
|  | 45-54 | 1 (1,1%) | 4 (3,9%) | 12 (7,1%) | 13 (4,1%) |  |
|  | 55-64 | 2 (1,7%) | 11 (4,1%) | 33 (10,1%) | 37 (8,1%) |  |
|  | 65-74 | 17 (15,5%) | 28 (7,8%) | 59 (10,8%) | 77 (10%) |  |
|  | 75+ | 30 (21,4%) | 84 (12,7%) | 187 (19,6%) | 185 (13,9%) |  |
| Death by BMI | <=20 | 1 (5,3%) | 3 (3,4%) | 20 (11,5%) | 24 (11,7%) | 9,39E-02 |
|  | 20-25 | 12 (9,2%) | 17 (5,6%) | 50 (11,1%) | 61 (6,7%) |  |
|  | 25-30 | 7 (3,6%) | 20 (5,2%) | 30 (7,2%) | 54 (5,5%) |  |
|  | 30< | 9 (5,2%) | 14 (4,1%) | 24 (7,2%) | 27 (3,4%) |  |

Overall in-hospital mortality was similar between COVID-19 (8%), influenza (8,1%) and other (8,6%) group, and markedly higher for the SARI group (13,5%). To limit the selection bias introduced by the current pandemic situation that would result in a more permissive hospitalization policy for the COVID-19 patients, we excluded mild COVID-19 cases. This analysis resulted in an in-hospital death rate of 12,6% for moderate to severe COVID-19 patients. When focusing on age groups 65-74 years and 75+ years, mortality rates of COVID-19 patients were 15,5% and 21,4%, respectively. Moreover, in-hospital mortality stratified by BMI showed that a lower proportion of patient with BMI over 30 *kg/m*^2^ died compared to the group with BMI between 20 *kg/m*^2^ and 25 *kg/m*^2^ (5,2 % and 9,2%, respectively).

### Comorbidities

As shown in Table 2 and depicted in Figure 2, the incidence rates for almost all analyzed comorbidities were lower among moderate to severe COVID-19 patients compared to other patients. A total of 9,5% of COVID-19 patients had cancer, compared to 17,1% of influenza patients and 23,4% of SARI patients. Accordingly, the age-adjusted odd ratio (AOR) for COVID-19 were 0,4 (95%CI: 0,27;0,59). Similar results were obtained when separating to solid-type cancer and hematologic cancer. In total, 3,5% of moderate to severe COVID-19 patients had chronic obstructive pulmonary disease (COPD), compared to 10%-11% of influenza and SARI patients and the AOR for COVID-19 was 0,29 (95%CI: 0,16;0,53). A total of 41,7% of COVID-19 patients had hypertension, compared to 52,6% of influenza patients and 47,6% of SARI patients, and the AOR for COVID-19 was 0,66 (95%CI: 0,5;0,87). A total of 52% of COVID-19 patients had cardiovascular disease, compared to 62%-64% of influenza and SARI patients. The AOR for COVID-19 was 0,46 (95%CI: 0,34;0,61). A total of 8,8% of COVID-19 patients smoked, compared to 20%-21% of influenza and SARI patients. The AOR for COVID-19 was 0,32 (95%CI: 0,22;0,48). Only dementia was more frequent among COVID-19 compared to other patients. A total of 10,1% of COVID-19 patients had dementia, compared to 4,9% of influenza patients and 8,2% of SARI patients. The AOR for COVID-19 was 2,82 (95%CI: 1,67;4,76). However, an interaction effect between dementia and arrival from nursing home was observed (p-value=0,0047). Stratified AOR led to nearly significant levels due to smaller sample size. COVID-19 patients hospitalized from nursing homes depicted an AOR of 1,82 (p-value=0,058), while the AOR for COVID-19 patients not coming from nursing homes was 0,54 (p-value=0,079).

**Table 2.** Comorbidities prevalence and age-adjusted odds ratios among study patients at admission and comparison between disease groups.

|  | COVID-19 *<br>(n=398) | Influenza<br>(n=1612) | SARI<br>(n=2292) | Others<br>(n=4054) | age-adjusted odds ratio (95%CI)<br>(COVID-19 vs. influenza/SARI) | FDR-corrected p-value<br>(odds ratio) |
| --- | --- | --- | --- | --- | --- | --- |
| Any comorbidity | 264 (66,3%) | 1316 (81,6%) | 1877 (81,9%) | 2766 (68,2%) | 0,32 (0,24;0,44) | 8,32E-12 |
| Smoking | 35 (8,8%) | 341 (21,2%) | 458 (20,0%) | 529 (13,0%) | 0,32 (0,22;0,48) | 2,05E-07 |
| Cardiovascular disease | 207 (52,0%) | 1037 (64,3%) | 1420 (62,0%) | 2169 (53,5%) | 0,46 (0,34;0,61) | 9,26E-07 |
| Any cancer | 38 (9,5%) | 276 (17,1%) | 536 (23,4%) | 731 (18,0%) | 0,4 (0,27;0,58) | 1,11E-05 |
| Chronic obstructive pulmonary disease (COPD) | 14 (3,5%) | 166 (10,3%) | 267 (11,6%) | 185 (4,6%) | 0,29 (0,16;0,53) | 0,0002 |
| Dementia | 40 (10,1%) | 79 (4,9%) | 188 (8,2%) | 224 (5,5%) | 2,82 (1,67;4,76) | 0,0003 |
| Solid cancer | 34 (8,5%) | 234 (14,5%) | 415 (18,1%) | 631 (15,6%) | 0,5 (0,33;0,75) | 0,0019 |
| Hematologic cancer | 4 (1,0%) | 51 (3,2%) | 140 (6,1%) | 123 (3,0%) | 0,19 (0,07;0,53) | 0,0028 |
| Hypertension | 166 (41,7%) | 848 (52,6%) | 1091 (47,6%) | 1699 (41,9%) | 0,66 (0,5;0,87) | 0,0064 |
| Ischemic heart disease | 57 (14,3%) | 358 (22,2%) | 452 (19,7%) | 664 (16,4%) | 0,69 (0,49;0,97) | 0,0583 |
| Asthma and bronchiectasis | 14 (3,5%) | 114 (7,1%) | 110 (4,8%) | 114 (2,8%) | 0,54 (0,28;1,03) | 0,0963 |
| Diabetes | 117 (29,4%) | 542 (33,6%) | 701 (30,6%) | 1031 (25,4%) | 0,78 (0,59;1,03) | 0,1159 |
| Stroke | 30 (7,5%) | 152 (9,4%) | 244 (10,6%) | 505 (12,5%) | 0,7 (0,42;1,16) | 0,2162 |
| Hyperlipidemia | 146 (36,7%) | 671 (41,6%) | 868 (37,9%) | 1305 (32,2%) | 0,87 (0,67;1,14) | 0,3333 |
| Osteoarthritis | 6 (1,5%) | 27 (1,7%) | 19 (0,8%) | 77 (1,9%) | 0,46 (0,1;2,02) | 0,3333 |
| Rheumatoid arthritis | 2 (0,5%) | 29 (1,8%) | 35 (1,5%) | 41 (1,0%) | 0,34 (0,04;2,61) | 0,3333 |
| Chronic kidney disease (CKD) | 51 (12,8%) | 256 (15,9%) | 281 (12,3%) | 464 (11,4%) | 0,92 (0,63;1,34) | 0,6721 |

\* excluding mild severity cases

**Fig. 2.**
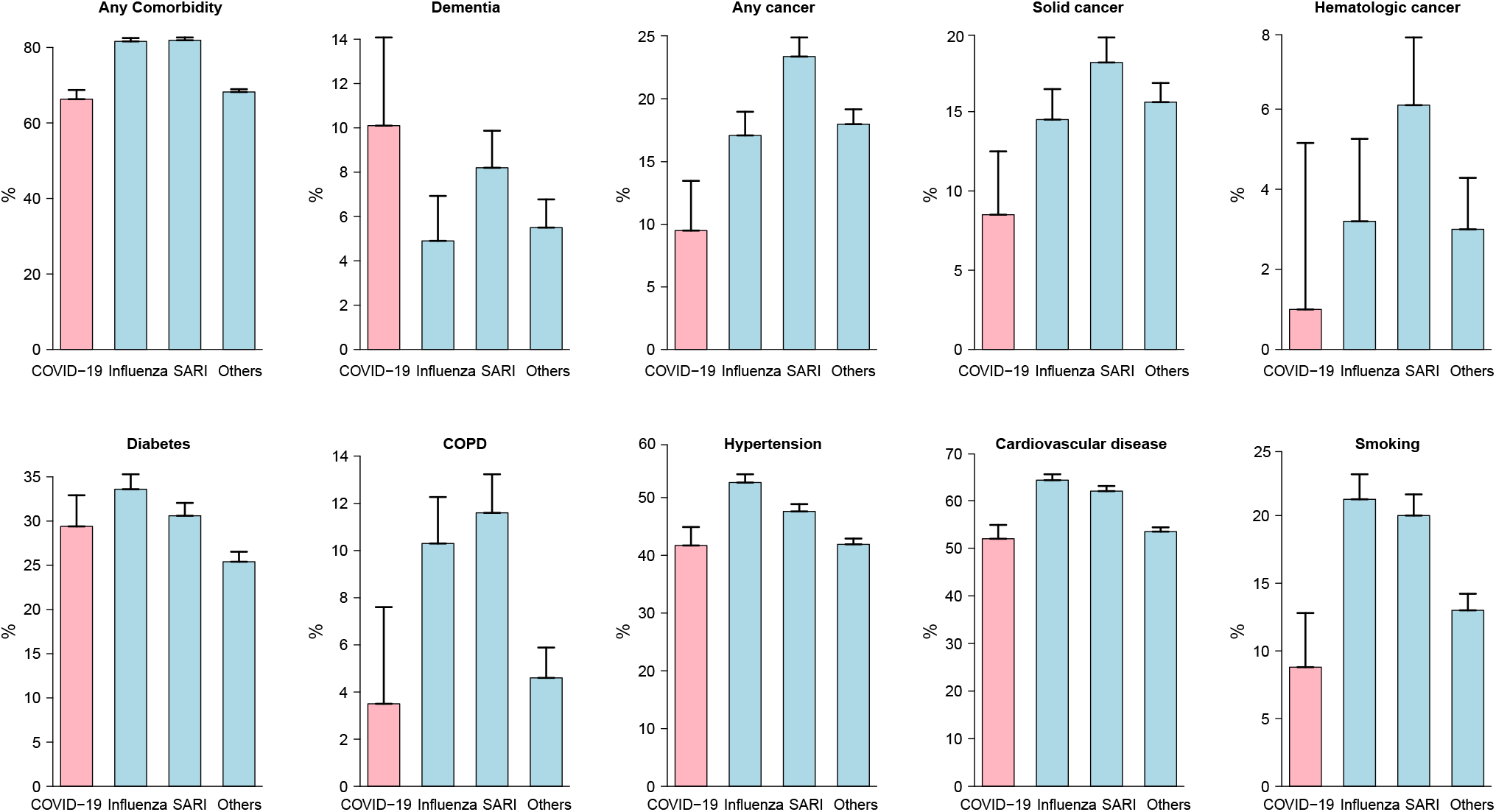
Comorbidity prevalence rates among study patients at admission. Moderate to severe cases were considered for COVID-19 patients. For all disease types but dementia, rates were lower among COVID-19 patients compared to other patients. The differences between disease groups were all statistically significant except for diabetes.

Lastly, diabetes was an important risk factor for COVID-19 severity, with a prevalence of nearly 30% of COVID-19 cases. Influenza and SARI depicted a similar prevalence suggesting diabetes as a common comorbidity of ILIs.

### Admission results

Table 3 and Table 4 present results of the intercohort comparison of vital signs and laboratory examinations at admission. The median standardized scores (see Methods) for each group are presented per variable within the heatmap in Figure 3.

**Table 3.** Vital Signs at admission.

|  |  | COVID-19 |  | Influenza |  | SARI |  | Others |  | FDR-corrected p-value<br>(group difference) |
| --- | --- | --- | --- | --- | --- | --- | --- | --- | --- | --- |
| Pulse (*bpm*) | n | 693 |  | 1612 |  | 2292 |  | 4047 |  | 5,28E-79 |
|  | median (IQR) | 88 | (23) | 93 | (28) | 97 | (29) | 86 | (25) |  |
| Systolic blood pressure (*mmHg*) | n | 692 |  | 1610 |  | 2288 |  | 4040 |  | 3,12E-14 |
|  | median (IQR) | 128 | (27) | 135 | (37) | 129 | (36,3) | 130 | (35) |  |
| Diastolic blood pressure (*mmHg*) | n | 693 |  | 1611 |  | 2291 |  | 4046 |  | 1,31E-21 |
|  | median (IQR) | 77 | (14) | 73 | (19) | 72 | (17) | 75 | (17) |  |
| Average blood pressure (*mmHg*) | n | 693 |  | 1612 |  | 2292 |  | 4047 |  | 4,80E-12 |
|  | median (IQR) | 102,5 | (18) | 104,5 | (24,5) | 100,5 | (24) | 103,5 | (24,3) |  |
| Room saturation (%) | n | 672 |  | 1572 |  | 2204 |  | 3880 |  | 2,88E-105 |
|  | median (IQR) | 96 | (4) | 95 | (6) | 95 | (6) | 97 | (3) |  |
| Oxygen saturation (%) | n | 319 |  | 1010 |  | 1475 |  | 2147 |  | 1,40E-59 |
|  | median (IQR) | 95 | (3) | 95 | (4) | 96 | (4) | 98 | (4) |  |
| Fever ($^{\circ}C$) | n | 689 | | 1610 | | 2291 | | 4031 | | 4,09E-127 |
|  | median (IQR) | 37,1 | (0,9) | 37,1 | (1,1) | 37,3 | (1,4) | 36,8 | (0,6) |  |
| Breaths (*num/min*) | n | 560 |  | 298 |  | 465 |  | 1333 |  | 1,85E-16 |
|  | median (IQR) | 17 | (5) | 16 | (6) | 18 | (8) | 16 | (6) |  |

**Table 4.** Laboratory examination at admission.

|  |  | COVID-19 | Influenza | SARI | Others | FDR corrected p-value<br>(group difference) |
| --- | --- | --- | --- | --- | --- | --- |
| White blood count (WBC) ($\cdot 10^3/\mu L$) | n<br>median (IQR) | 657<br>6,5 (3,6) | 1600<br>8,2 (5,2) | 2262<br>11,3 (8,2) | 3960<br>10,4 (6) | 5,81E-153 |
| Platelets ($\cdot 10^3/\mu L$) | n<br>median (IQR) | 656<br>200 (100) | 1589<br>187 (97) | 2273<br>218 (131) | 3969<br>221 (112) | 1,15E-34 |
| Hemoglobin (g/dL) | n<br>median (IQR) | 657<br>13,2 (2,4) | 1604<br>12,4 (2,8) | 2290<br>11,9 (3) | 3986<br>12,3 (2,8) | 2,34E-34 |
| Lymphocytes (%) | n<br>median (IQR) | 633<br>19,9 (15,5) | 1587<br>11,5 (10,9) | 2250<br>10,4 (11) | 3637<br>13,9 (13,2) | 1,53E-74 |
| Lymphocytes (abs) ($\cdot 10^3/\mu L$) | n<br>median (IQR) | 633<br>1,2 (0,9) | 1557<br>0,9 (0,8) | 2166<br>1,1 (0,9) | 3603<br>1,4 (1,1) | 9,54E-86 |
| Neutrophils (%) | n<br>median (IQR) | 633<br>69,6 (18,2) | 1589<br>77,5 (15,1) | 2280<br>78,4 (15,3) | 3688<br>75,6 (16,9) | 8,44E-27 |
| Neutrophils (abs) ($\cdot 10^3/\mu L$) | n<br>median (IQR) | 633<br>4,4 (3,1) | 1558<br>6,1 (4,4) | 2182<br>8 (6,2) | 3639<br>7,5 (5,4) | 9,56E-78 |
| Creatinine (mg/dL) | n<br>median (IQR) | 658<br>0,9 (0,4) | 1593<br>1 (0,6) | 2284<br>1 (0,7) | 3698<br>0,9 (0,6) | 6,62E-08 |
| Blood urea nitrogen (BUN) (mg/dL) | n<br>median (IQR) | 658<br>14,2 (10,3) | 1592<br>18,9 (14) | 2280<br>19 (16,6) | 3686<br>17,5 (14,8) | 6,37E-25 |
| Potassium (mEq/L) | n<br>median (IQR) | 658<br>3,9 (0,6) | 1591<br>4 (0,6) | 2274<br>4 (0,7) | 3749<br>4,1 (0,6) | 9,67E-13 |
| Sodium (mEq/L) | n<br>median (IQR) | 660<br>136 (5) | 1594<br>135 (5) | 2269<br>135 (6) | 3742<br>137 (5) | 1,36E-29 |
| D-dimer (mg/L) | n<br>median (IQR) | 462<br>701,5 (751,8) | 83<br>3,9 (827,2) | 107<br>929 (1767) | 178<br>1268,5 (1500,8) | 2,20E-20 |
| Ferritin (ng/mL) | n<br>median (IQR) | 11<br>377 (598) | 209<br>212 (387) | 225<br>343 (746) | 441<br>224 (514) | 2,20E-05 |
| C-reactive protein (CRP) (mg/L) | n<br>median (IQR) | 622<br>4,7 (10,4) | 685<br>7,3 (16,1) | 904<br>13,9 (19,2) | 2394<br>5,6 (12,7) | 1,75919E-70 |
| Procalcitonin (ng/mL) | n<br>median (IQR) | 38<br>0,4 (0,7) | 14<br>0,7 (2,3) | 36<br>0,5 (1,2) | 85<br>0,4 (1,3) | 2,23E-01 |
| Troponine (ng/mL) | n<br>median (IQR) | 67<br>14 (40,5) | 462<br>12 (44,8) | 421<br>21 (71) | 782<br>33 (129,8) | 9,57E-26 |
| Brain natriuretic peptide (BNP) (pg/mL) | n<br>median (IQR) | 78<br>446,9 (2851,9) | 164<br>2323,4 (4753) | 204<br>2700 (5891,9) | 559<br>2232,1 (5991,5) | 1,30E-06 |
| Lactate dehydrogenase (LDH) (U/L) | n<br>median (IQR) | 451<br>258 (155,5) | 431<br>226 (134) | 550<br>232,5 (141,3) | 1132<br>230 (140,8) | 2,20E-04 |
| Aspartate aminotransferase (AST) (U/L) | n<br>median (IQR) | 622<br>31 (22) | 1386<br>31 (24) | 1812<br>27 (24) | 3058<br>26 (23) | 2,60E-14 |
| Alanine aminotransferase (ALT) (U/L) | n<br>median (IQR) | 473<br>22 (23) | 1267<br>26 (23) | 1520<br>23 (26) | 2356<br>20 (24) | 3,02E-17 |

**Table 4.bis.** Laboratory examination at admission.

|  |  | COVID-19 |  | Influenza |  | SARI |  | Others |  | FDR corrected p-value<br>(group difference) |
| --- | --- | --- | --- | --- | --- | --- | --- | --- | --- | --- |
| International normalized ratio (INR) | n | 523 |  | 993 |  | 1331 |  | 2336 |  | 1,23E-18 |
|  | median (IQR) | 1,1 | (0,1) | 1 | (0,2) | 1,1 | (0,2) | 1,1 | (0,2) |  |
| Prothrombin time (PT) (sec.) | n | 523 |  | 994 |  | 1331 |  | 2336 |  | 7,19E-33 |
|  | median (IQR) | 12,9 | (1,7) | 11,9 | (2,5) | 12,6 | (2,9) | 12,7 | (2,5) |  |
| Partial thromboplastin time (PTT) (sec.) | n | 522 |  | 987 |  | 1327 |  | 2319 |  | 3,46E-34 |
|  | median (IQR) | 31,9 | (4,8) | 29,1 | (6,6) | 29,4 | (6,2) | 30,3 | (6,2) |  |
| Albumin (g/dL) | n | 461 |  | 1302 |  | 1641 |  | 2752 |  | 4,09E-127 |
|  | median (IQR) | 3,8 | (0,6) | 3,4 | (0,8) | 3,1 | (0,8) | 3,4 | (0,8) |  |
| Bilirubin (mg/dL) | n | 632 |  | 1402 |  | 1834 |  | 3113 |  | 7,15E-16 |
|  | median (IQR) | 0,5 | (0,3) | 0,5 | (0,4) | 0,5 | (0,5) | 0,5 | (0,5) |  |
| Creatine phosphokinase (CPK) (U/L) | n | 491 |  | 394 |  | 490 |  | 1301 |  | 1,83E-04 |
|  | median (IQR) | 85 | (118,5) | 90,5 | (202) | 73,5 | (134,5) | 84 | (189) |  |
| Glucose (mg/dL) | n | 660 |  | 1596 |  | 2282 |  | 3785 |  | 1,36E-36 |
|  | median (IQR) | 107 | (43) | 123 | (63) | 129 | (70) | 119 | (61) |  |
| Calcium (blood) (mg/dL) | n | 636 |  | 1482 |  | 2061 |  | 3428 |  | 5,61E-11 |
|  | median (IQR) | 8,8 | (0,9) | 8,6 | (0,8) | 8,6 | (0,9) | 8,7 | (1) |  |
| Calcium (ionized) (mg/dL) | n | 566 |  | 1331 |  | 1920 |  | 3064 |  | 1,37E-86 |
|  | median (IQR) | 1,2 | (0,1) | 1,1 | (0,1) | 1,2 | (0,1) | 1,2 | (0,1) |  |
| Magnesium (mg/dL) | n | 498 |  | 1386 |  | 1896 |  | 2928 |  | 2,84E-44 |
|  | median (IQR) | 2,1 | (0,3) | 1,9 | (0,4) | 1,9 | (0,4) | 2 | (0,3) |  |
| Phosphorus (mg/dL) | n | 503 |  | 1387 |  | 1897 |  | 2947 |  | 5,85E-11 |
|  | median (IQR) | 3,1 | (1) | 3,3 | (1,2) | 3,3 | (1,2) | 3,4 | (1,2) |  |
| Fibrinogen (mg/dL) | n | 472 |  | 202 |  | 336 |  | 959 |  | 3,79E-18 |
|  | median (IQR) | 413,5 | (152) | 391,6 | (191,2) | 459 | (270,5) | 388 | (192) |  |
| PCO2 (mmHg) | n | 566 |  | 1333 |  | 1924 |  | 3066 |  | 4,07E-09 |
|  | median (IQR) | 44 | (11) | 47 | (14) | 46 | (14) | 46 | (14) |  |
| Bicarbonate (mmol/L) | n | 566 |  | 1331 |  | 1920 |  | 3059 |  | 2,04E-33 |
|  | median (IQR) | 26,6 | (4,3) | 27,9 | (6,2) | 27,8 | (6,3) | 26 | (6,3) |  |
| pH | n | 566 |  | 1333 |  | 1924 |  | 3066 |  | 4,28E-30 |
|  | median (IQR) | 7,4 | (0,1) | 7,4 | (0,1) | 7,4 | (0,1) | 7,4 | (0,1) |  |
| Lactate (mmol/L) | n | 287 |  | 384 |  | 674 |  | 1256 |  | 2,09E-14 |
|  | median (IQR) | 1,3 | (0,9) | 1,7 | (1,5) | 1,8 | (1,8) | 1,7 | (2) |  |
| MetHemoglobin (g/dL) | n | 121 |  | 13 |  | 29 |  | 47 |  | 1,68E-04 |
|  | median (IQR) | 0,5 | (0,5) | 1 | (0,8) | 1 | (0,4) | 1,1 | (0,9) |  |
| CarboxiHemoglobin (g/dL) | n | 140 |  | 12 |  | 27 |  | 42 |  | 8,63E-05 |
|  | median (IQR) | 1,4 | (1) | 1,9 | (0,9) | 1,9 | (0,9) | 1,6 | (0,8) |  |

**Fig. 3.**
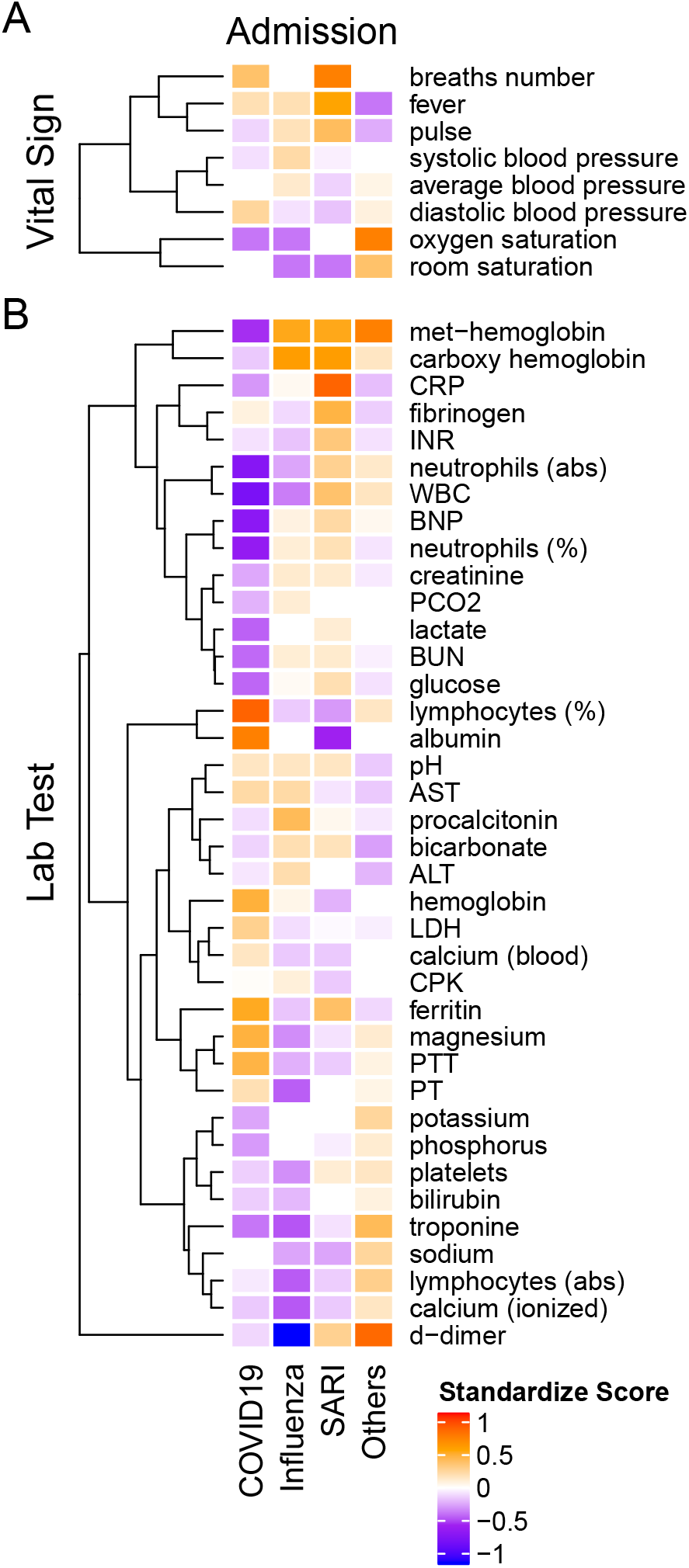
Heatmap of admission median standardized scores of (A) vital signs and (B) lab tests in COVID-19, influenza A/B, SARI and Others. All variables are introduced on the same scale, relative to the variable overall median. The standardized scores is positive if the group median is greater than the overall median (orange-red) and negative otherwise (purple-blue). Hierarchical clustering on the left points to groups with similar pattern across diseases.

#### Vital signs at admission

The median standardized score (Figure 3A) of the heart rate (pulse) among COVID-19 patients (median 88 ± 23 IQR *bpm*) was substantially lower compared to influenza (median 93 ± 28 IQR *bpm*) and SARI (median 97 ± 29 IQR *bpm*) patients. Systolic blood pressure was lower among COVID-19 patients (median 128 ±27 *mmHg*) compared to influenza patients (median 135± 37 *mmHg*), while diastolic blood pressure was higher among COVID-19 patients (median 77 ± 14 *mmHg*) compared to influenza (median 73 ± 19 *mmHg*) and SARI patients (median 72 ± 17 *mmHg*). The standardized scores (in Figure 3) showed that among the vital signs, blood pressure parameters (systolic, diastolic and average) were regrouped as a single cluster. Similarly, saturation parameters (oxygen and room) formed a single group. A third group included pulse, fever and breath number. However, neither respiratory measures nor saturation or temperature showed clinically relevant median differences between ILIs.

#### Lab tests at admission

Laboratory examinations are presented in Table 4 and in Figure 3B. Lymphocytes percentage among COVID-19 patients (median 19,9 ± 15,5 IQR %) was substantially higher compared to influenza patients (median 11,5 ± 10,9 IQR %) and SARI patients (median 10,4 ± 11 IQR %). Similarly, albumin levels were higher among COVID-19 patients (median 3,8 ± 0,6 IQR *g/dL*) compared to influenza patients (median 3,4 ± 0,8 IQR *g/dL*) and SARI patients (median 3,1± 0,8 IQR *g/dL*). The standardized scores showed that among the lab test results, magnesium, prothrombin time (PT), partial thromboplastin time (PTT), hemoglobin, lactic dehydrogenase (LDH) and calcium, as well as of lymphocytes percentage, were regrouped as a single cluster that indicated higher levels among COVID-19 patients as compared to influenza and SARI. Conversely, neutrophils (abs), White blood count (WBC), brain natriuretic peptide (BNP) and neutrophils percentage formed a single cluster that indicated substantially lower levels in COVID-19 patients as compared to influenza and SARI. A similar effect with reduced levels in COVID-19 patients as compared to influenza and SARI was shown for the cluster containing methemoglobin and carboxyhemoglobin, the cluster containing procalcitonin, bicarbonate, alanine aminotransterrase (ALT), potassium and phosphorous, and the cluster containing creatinine, lactate, blood urea nitrogen (BUN), and glucose.

#### Disease trajectories

Trends over time of parameters that showed differences between patient groups, are presented in Table 5. The results for of all pairwise group comparisons are provided as supplementary Table S2. For each variable and compared pair, the table provides the time gap for which the stronger difference in rank means slopes was found, either (0-6, 6-24) or (6-24, 24-48), and the difference itself. Figure 4 represents the trends for each of the variables showing a significant effect on diseases trajectories. As shown by the trends in Figure 4, both systolic and diastolic blood pressures, as well as pulse, decreased more slowly during the 6-48 hours after admission among COVID-19 patients compared to influenza and SARI patients. The difference in slopes with respect to its significance level (p-value) are represented in the volcano plot in Figure 5. The left panel refers to all influenza-COVID-19 comparisons, and the right panel refers to all SARI-COVID-19 comparisons. COVID-19 patients depicted a slower decrease in fever during the 48 hours after admission, compared to SARI patients. A particularly distinctive behavior among COVID-19 patients was noted for variables relating to WBC, which slightly increased among COVID-19 patients during the 24 hours after admission, but declined in all other patients. COVID-19 patients, similarly to SARI patients, first showed a decrease, and then an increase in lymphocytes percentage during the first 48 hours after admission, while influenza patients showed a persistent increase. Neutrophils percentage and count also showed a difference in trend across time between COVID-19 patients and influenza patients. In particular, COVID-19 patients showed a consistent increase in percentage, compared to the decrease observed in patients of other cohorts. Finally, glucose levels among COVID-19 patients were relatively stable during the first 24 hours after admission, and then increased, as opposed to all other groups, where a consistent decrease was observed during the 48 hours after admission.

**Table 5.** Significant slope difference in time window in the first day or the second day of hospitalization.

| variable | disease compared to COVID-19 | time gap of max. difference | FDR corrected p-value |
| --- | --- | --- | --- |
| systolic blood pressure | influenza | (0-6)-(6-24) | 4,30E-06 |
| average blood pressure | influenza | (6-24)-(24-48) | 5,22E-06 |
| diastolic blood pressure | SARI | (0-6)-(6-24) | 6,54E-05 |
| fever | SARI | (0-6)-(6-24) | 0,0001 |
| diastolic blood pressure | influenza | (6-24)-(24-48) | 0,0019 |
| neutrophils (abs) | influenza | (0-6)-(6-24) | 0,0040 |
| average blood pressure | SARI | (0-6)-(6-24) | 0,0056 |
| lymphocytes (%) | influenza | (0-6)-(6-24) | 0,0057 |
| SAO<sub>2</sub> | SARI | (0-6)-(6-24) | 0,0109 |
| pulse | influenza | (6-24)-(24-48) | 0,0124 |
| glucose | influenza | (6-24)-(24-48) | 0,0141 |
| glucose | SARI | (6-24)-(24-48) | 0,0152 |
| WBC | SARI | (6-24)-(24-48) | 0,0220 |
| neutrophils (%) | influenza | (0-6)-(6-24) | 0,0232 |
| WBC | influenza | (0-6)-(6-24) | 0,0242 |

**Fig. 4.**
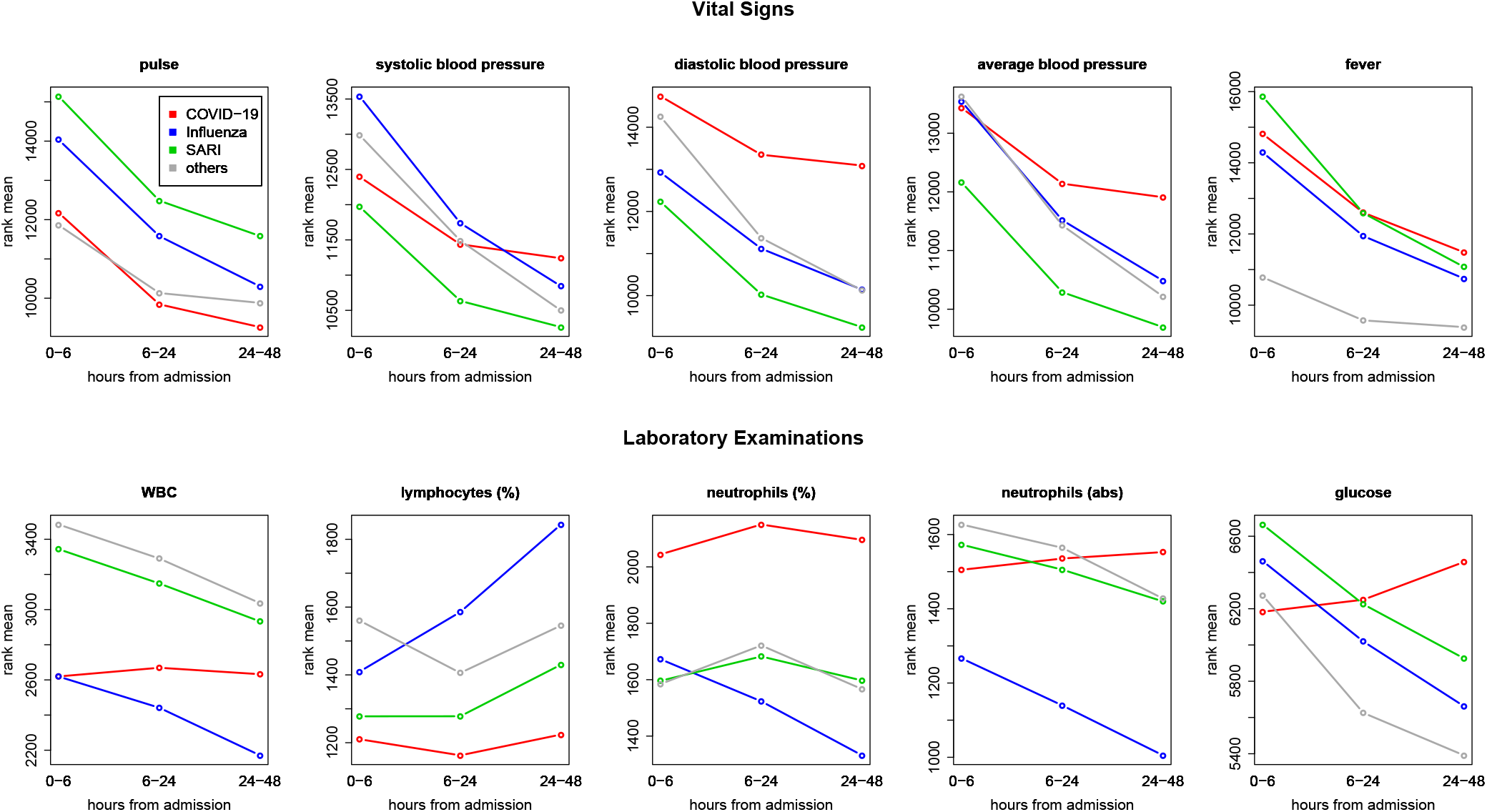
Comparing trends across time – ranks means vs. time intervals. Blood pressure and pulse were decreasing more slowly during 6-48 hours after admission among COVID-19 patients compared to influenza and SARI patients. Glucose and measures related to white blood cells (white blood count, lymphocytes and neutrophils) among COVID-19 patients showed distinct trajectories, with respect to other patients.

**Fig. 5.**
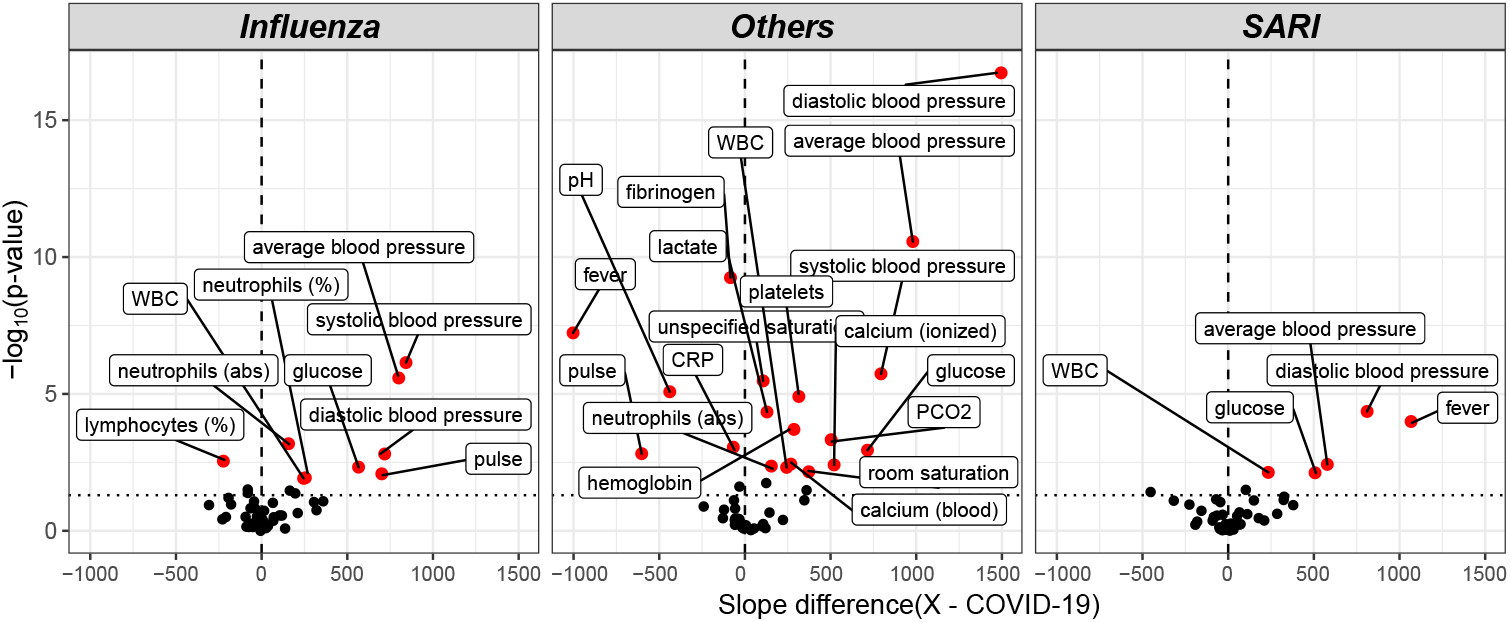
Volcano plot of pairwise post-hoc analysis of patients trajectory of COVID-19, influenza, SARI and others for each lab tests. The level of significance is shown vs. the effect size, namely the slope difference (x - COVID-19). The most highly significant result was for average blood pressure, as seen at the left window, which refers to all influenza-COVID-19 comparisons, and at the right window, which refers to all SARI-COVID-19 comparisons. Trends for WBC, glucose and diastolic blood pressure for COVID-19 patients were found to be different from both influenza and SARI.

## Discussion

The relative virulence of COVID-19 versus influenza has been and is still debated. Therefore, our focus was to assess COVID-19 mortality and virulence compared to other ILIs. In-hospital death rates were investigated. While COVID-19 depicted a lower overall mortality, stratification by age showed an increased fraction of casualty among elderly persons consistent with other reports (24). Age has been associated with COVID-19 severity and mortality in plethora of studies (1, 2, 4–7, 9). The presented comorbidity analysis exposed striking differences between disease groups. COVID-19 patients depicted substantially fewer “overall comorbidities” than influenza or SARI patients (66.3% versus 81.6% and 81.9%). The difference remained after age adjustment and excluding mild severity, suggesting that COVID-19 patients required hospitalization more frequently even if they were healthier before their infection. Hypertension, cardiovascular disease and diabetes depicted a large prevalence in COVID-19, similarly to previous findings (3, 6, 25), and represented common risk factors for ILIs. However, only hypertension and cardiovascular disease showed a significantly lower prevalence in COVID-19 patients compared to other ILIs. Interestingly, cancer prevalence was lower among COVID-19 patients, although it remains unclear how different cancers and/or cancer therapies affect COVID-19 severity or SARS-CoV2 infectivity (26). A recent study suggested that the differential immune cell profiles of cancer patients treated with immunomodulatory agents, may impact the host response to the SARS-COV2 diminish disease severity (27).The lower prevalence can also be explained by higher self COVID-19 risk perception among cancer patients that leads to higher adherence to isolation methods. Taken together, the present investigation demonstrates that COVID-19 hospitalized patients present a significantly lower amount of comorbidities and are younger. COVID-19 patients hospitalized with moderate to severe disease had a significantly higher mortality rate than hospitalized influenza patients. Overall, this suggests that the intrinsic virulence of COVID-19 is higher than influenza.

Our analysis of vital signs revealed that pulse, blood pressure and temperature (fever) were significantly different between ILIs at admission and showed different patterns in the two days post admission despite the small effect size on fever. An independent study on acute respiratory syndrome (10) comparing Influenza and COVID-19 patients (severe) showed that partial pressure of oxygen in arterial blood (*P aO*_2_) was remarkably low for COVID-19 patients compared to influenza. Hence, oxygen saturation, heartbeat, temperature, and others continuous measures (i.e. blood glucose, activity) may serve as informative parameters to be continuously monitored in COVID-19 patients. For example, such measurements could be used to anticipate hospitalization, at the point of care, or to predict potential readmission (28–35). Monitoring vital signs at the hospital is critical for following patient disease trajectory (36) and for guiding physicians on clinical interventions such as initiation of ventilation assistance or pharmaceutical treatment.

Obesity has been reported as a major risk factor for COVID-19 and type 2 diabetes. Furthermore, impaired metabolic health (i.e., dyslipidaemia and insulin resistance) is linked with a higher risk of COVID-19-associated pneumonia (37–39). The presented analysis showed that BMI was indeed significantly higher in COVID-19 patients with respect to SARI and Influenza, while diabetes prevalence was high but similar to other ILIs (∼30%). The lower BMI group depicted a higher mortality as compared to obese patients. However, due to the high proportion of missing data on BMI among the patients that died and the relatively small sample size, the observed differences warrant further investigations. This obesity paradox has been reported elsewhere (40) and seems to be common between ILIs (41). Several hypothesis have been proposed to explain why obese patients are more affected by COVID-19 without an increased mortality (42). For instance, Adipocytes contains the ACE2 receptor that enable the entrance of the COVID-19 virus, which turns adipose tissue into a potential target and viral reservoir (43). On the other side, Influenza infection relies on a different mechanism, through hemagglutinin onto sialic acid sugars on the surfaces of epithelial cells (44) which could explain a higher severity of COVID-19 disease in overweight patients. Conversely, in obese patients, an increased circulating levels of adipokines and inflammatory cytokines such as TNFα, IL-6, or C-Reactive Protein (CRP) was observed (45). This chronic low-grade inflammation may impair the adaptive immune responses to viral infections (46) and consequently reduce the probability of a lethal cytokine storm. However, it remains controversial for the COVID-19 disease and the underlying mechanisms remains unclear (47).

At the laboratory examination level, several metabolic intermediates and enzymes were significantly different between COVID-19, SARI, and influenza patients, particularly, lactate, LDH and glucose. Intriguingly, glucose levels in COVID-19 patients were lower at admission but took an opposite trajectory in the first two days of admission, compared to SARI and influenza. Impaired glucose homeostasis is associated with poor COVID-19 prognosis and has been hypothesized as an underlying trigger of the cytokine storm in COVID-19 patients (48–51). In severe cases of COVID-19, a hyperinflammatory response (cytokine storm) is correlated with poor outcome (52–54). Several studies reported an association between the neutrophils-to-lymphocyte ratio (NLR) and the severity of COVID-19 (55–57). Accordingly, our investigation of lab examination revealed that COVID-19 have a prominent effect on blood cell-types counts and proportions such as lymphocytes, neutrophils, WBCs at admission and along the first two days of hospitalization. The present analysis reveal that the trajectories of glucose and immune cells are affected by the clinical management of COVID-19 patients, probably through corticosteroid treatments (58).

Ferritin is a key player of immune dysregulation through its immune-suppressive and pro-inflammatory effects, supporting the possibility of cytokine storm (59, 60). We observed that ferritin levels were more elevated among COVID-19 patients as compared to SARI or influenza patients at admission. However, here the ferritin test was performed only for a small number (∼ 1, 5%) of severe COVID-19 patients.

A non-neglectable risk of thrombosis or disseminated intravascular clot formation has been described for COVID-19 patients (53, 61). Histopathological studies in post-mortem lung tissue revealed pulmonary microthrombi in 57% of COVID-19 and 58% of SARS as compared to 24% of H1N1 influenza patients (62). However, the underlying mechanisms remain to be fully characterized. It has been hypothesized that the dysregulated immune responses orchestrated by inflammatory cytokines is involved. Interestingly, our analysis of laboratory tests at admission confirmed higher level of PT and PTT in COVID-19 patients while platelet levels did not show extreme values for COVID patients with respect to influenza or SARI patients. In addition, D-dimers depicted a much lower value in influenza compared to COVID-19 or SARI patients. However, this observation might be biased due to the low number of tests made for influenza patients (∼5%). As the risk due to thrombosis has been described early in the pandemic (63), D-dimer quantification is currently performed routinely for COVID-19 patients.

The diagnosis of dementia has been reported as an important risk factor for mortality in COVID-19 patients (64–66). The prevalence of demented patient found in the COVID-19 cohort (∼10%) was lower than previous estimates, which vary from 13% to 42% (67). According to our analysis, dementia was found to be more prevalent among COVID-19 patients as compared to patients of the other cohorts. In order to determine if this finding can be explained by the higher proportion of COVID-19 patients arriving from nursing homes, the variable of nursing home residence was added to the model. A significant interaction between the nursing home residence variable and dementia in COVID-19 patients was observed.

The results can be potentially explained by a higher exposure and infection risk of demented nursing home residents caused by a lower ability to adhere to the needed isolation behaviors (68). COVID-19 exposure in nursing home has been reported by others and is a common issue in several countries (24, 69). More than 20% of all reported COVID-19-associated deaths occurring in nursing homes in countries such as Canada, Sweden and the UK. For instance, nursing home population depicted a 1.70-fold higher infection attack rate than the general population in France (24, 69). Thus, COVID-19 exposure in nursing home has been reported by others and is a common issue in several countries.

The above findings point to vital signs and lab results, as well as to comorbidities, that can be helpful in discriminating between COVID-19 and other ILI’s at presentation. We plan to pursue a study to develop data-driven tools to efficiently identify COVID-19 cases and predict their disease trajectory at the point of care.

Haifa and the Northern district of Israel are a multicultural region that encompasses several ethnic groups, who have distinct cultural agendas and socio-economic characteristics. The data collected from the largest regional hospital provided were of value for assessment of differential COVID-19 manifestations and risk factors in various ethnic groups. A striking enrichment of all COVID-19 cases in Arab communities accounting for about 40% of COVID-19 cases was observed, as opposed to roughly 25% for the influenza and SARI cases, similar to the Arab proportion in the Haifa area. Higher exposure risk and infection rates of COVID-19 at Arab settlements may account for this effect as no differences in terms of severity between Arabs and Jews were observed. Further stratification of the Jewish population could not be performed for a more detailed analysis of the orthodox or secular fraction. However, it was previously reported that a similar effect is likely in the orthodox Jewish communities (70–72).

Other studies that compared COVID-19 to Influenza did not refer to associations between variables, which may be attributed to systematic activity, and to their clinical trajectory, and used limited cohorts of patients or focused only on acute respiratory distress syndrome, for instance (10, 11). Furthermore, they did not account for multiple testing of variables, which may lead to increased overall type I error (19). In this study, a comprehensive approach was applied to accommodate simultaneous testing of multiple biomarkers. Heterogeneity in distribution between variables was addressed by unifying their scale through robust standardization, thereby allowing comparison of their effects and detection of common patterns through clustering. Non-parametric repeated measures model was performed to allow for time-course analysis under various forms of non-symmetric, long-tailed distributions.

### Study limitations

The present study has few important limitations. Firstly, this study is based on a single medical center. Secondly this analysis did not include important information such as symptoms, images (scans, x-rays), waveforms (e.g. electrocardiogram, oxygen measurements in blood, respiration, ventilation), omics data (e.g. genomics, epi-genomics, transcriptomics, lipidomics and metabolomics) and pharmacological interventions (apart from the severe cases definition). We decided not to include oxygen measurement because of the uncertainly as per whether the measurement was taken on an arterial or venous line. Thirdly, potential selection bias, information bias or non differential misclassification associated with the use of EMR data are existing in the presented dataset. Variables were collected from a unique EMR sources and using a single query tools. These EMR were curated similarly for each groups which allowed to observe clinically relevant significant differences. Differences in the prevalence of personal characteristics between the diseases among hospitalized patients can be related to differences in exposure risk, infection susceptibility, disease severity and admission to hospital policies and biases. To address the effect of selection bias related to differences in hospitalization admission policy of COVID-19 patients compared to the other diseases, asymptomatic and mild cases were excluded in part of the analysis. Asymptomatic and mild cases are usually not hospitalized in influenza and SARI groups unless combined with other risk factors (Berkson’s bias)(73). Furthermore, some of the lab tests are not part of the routine work, hence selection bias by indication is likely. Not all patients within the SARI group were tested for Influenza, COVID-19 or other viruses, thus the SARI group potentially contains undetected Influenza cases, and less likely, COVID-19 cases, as the vast majority patients with Influenza or SARI in our cohort were diagnosed prior to COVID-19 emergence. Nevertheless, the SARI group is heterogeneous in diagnoses, while the Influenza group, which is the focus in this paper for comparison to the COVID-19 group, remains homogeneous and clearly defined. Similar potential misclassification of influenza cases within the “Others” group is expected to be negligible. This group contains cases with a negative COVID-19 test result, at a period starting at the emergence of COVID-19 in Israel, which was the end of the 2020 influenza season, and ending before the beginning of the next influenza season (74).

## Conclusions

The intrinsic virulence of COVID-19 appears higher than influenza. Several critical functions, such as immune response, coagulation, heart and respiratory function and metabolism were markedly impacted by the COVID-19 disease, despite some similarities observed with influenza and SARI. Moreover, COVID-19 seems to differently affect specific segments of the population, potentially due to increased exposure in localized communities or in nursing homes.

## Supporting information

Table S1 ICD9 codes

Table S2 Trajectory statistics

## Data Availability

The description of the tables and variables included in COV19 database is available on the online resource site. Future access to the ressource can be given to interested researchers, subject to hospital IRB approval.

https://cov19-resource.com/

## ACKNOWLEDGEMENTS

This research is partially supported by The Milner Foundation, founded by Yuri Milner and his wife Julia. We are grateful to the Placide Nicod fundation for their financial support (J.S.). We acknowledge the financial support of the Technion Machine Learning Intelligent Systems center.

## Notes

### Competing Interest Statement

The authors have declared no competing interest.

### Author Declarations

Ethical approval for this research was provided by the local institutional review board (IRB; #0141-20) Rambam Health Care Campus Haifa, Israel.

### Summary of Updates

-Improved discussion on dementia and COVID-19 -Clarifications on group definitions -Improved discussion on potential bias (classification bias)

